# Development and validation of a user-friendly smartphone imaging and telemedicine platform for remote diagnosis of anterior segment eye disease

**DOI:** 10.1101/2025.11.11.25339801

**Authors:** Kunal S. Parikh, Jordan Shuff, Kamini Reddy, Xiangrong Kong, Ximin Li, Shreya Hariharakumar, Vijay Antony Santhanaraj, Rashmirita Kakoty, Bharadwaj Karnam, Prabhu Krishna Ravilla, Nishant Vivekanand, Mary Hoopes, Kelsey Detels, Niam Mohseni, Rohan Verma, Dema Shumeyko, Dayakar Yadalla, Rengaraj Venkatesh, Nakul S. Shekhawat

## Abstract

**Background:** Cataract and anterior segment diseases are leading causes of blindness in low-resource settings. Eye camp screenings remain the primary mode of community outreach but are constrained by cost, logistics, and dependence on highly trained specialists. We designed and validated a low-cost, user-friendly smartphone-based anterior segment imaging and teleophthalmology platform to enable community health workers (CHWs) to perform diagnostic-quality eye screening.

**Methods:** We designed a portable imaging device (Scout^TM^) paired with an accessible Android smartphone and mobile application (InSightful) for telemedicine in low-bandwidth settings. CHWs underwent 3 hours of training on using the imaging software and hardware, then screened patients across 19 rural eye camps in South India with anterior segment images and clinical data uploaded to a cloud-based database for remote ophthalmologist (RO) review. Diagnoses and referral decisions made by ROs were compared with those of in-person eye camp ophthalmologists (ECOs). CHWs, ROs, and patients were surveyed on the platform’s feasibility and acceptability.

**Findings:** N=1093 patients underwent eye camp screening by ECOs and CHW-led smartphone screening with RO review. CHWs completed screenings in <2.5 minutes/eye and obtained diagnostic-quality images for >90% of eyes. ROs and ECOs showed 96.1% concordance for referral decisions (95% CI 94.7-97.1) and substantial agreement in diagnosis of any cataract (κ 0.77, percent agreement [PA] 89%), mature cataract (κ 0.67, PA 96%), immature cataract (κ 0.69, PA 85%), clear crystalline lens (κ 0.65, PA 89%), pseudophakia (κ 0.92, PA 97%), and moderate agreement for pterygium (κ 0.47, PA 94%). Concordance increased with image quality. CHWs, ROs, and patients reported high usability, acceptability, and net promoter scores.

**Interpretation:** Scout^TM^ anterior segment screening by minimally trained CHWs achieves diagnostic and referral accuracy comparable to in-person ophthalmologist examinations, supporting potential to decentralize cataract screening and expand access to eye care in low-resource settings.

**Funding:** National Eye Institute R21EY034343, National Eye Institute K23EY032988, National Eye Institute P30EY01765 (Biostatistics Core), Microsoft Innovation Acceleration Award, Johns Hopkins Center for Global Health, Stephen F Raab and Mariellen Brickley-Raab Rising Professorship in Ophthalmology, Boone Pickens Rising Professorship in Ophthalmology

## 1. INTRODUCTION

Anterior segment diseases such as cataract, corneal infection, corneal scarring, and pterygium are leading causes of vision loss and ocular morbidity, particularly in low-resource settings.^1,2^ Cataract accounts for 45% of global blindness, causing blindness or visual impairment in over 100 million people worldwide.^1,3^ Corneal infections and opacities cause blindness in up to 2 million people per year.^4^ However, image-based screening for anterior segment diseases has lagged behind posterior segment screening due to challenges with cost, portability, image quality, and usability of anterior segment imaging devices. Conventional tabletop slit lamps are expensive (typically $12,000-$17,000 USD),^5^ non-portable, and have a steep learning curve while portable systems lack standardized illumination, focus, magnification,^6^ or intuitive hardware and software interfaces. As a result, the majority of cataract and cornea screenings worldwide continue to depend on in-person ophthalmologist examinations despite an increasing burden of age-related eye diseases, increasing ophthalmic workforce shortages,^2,3^ persistent inequities in eye care access,^3,7–9^ and near-universal mobile and internet connectivity with steadily narrowing coverage gaps in rural regions.^10^

In low- and middle-income countries (LMICs), cataract and anterior segment screening largely relies on eye camps, which are periodic outreach events in which ophthalmologists and/or optometrists travel from urban hospitals to rural sites for in-person patient screening at predetermined dates, times, and locations.^11^ These programs are constrained by limited eye specialist availability, high cost, and logistical complexity, reaching only a fraction of rural populations. For example, an Aravind Eye Hospital (AEH) study found that eye camps screened only 7% of rural residents in a given region, with attendance dropping 80% for those living >3 km away.^12^ Of patients that did not attend, 44% were visually impaired and one-third needed cataract surgery.^12^ Barriers to access disproportionately affect vulnerable populations including women,^3,13^ older adults, and poor or rural patients.^3^ The COVID-19 pandemic further underscored these vulnerabilities as crowding restrictions halted outreach activities and severely reduced rural access to eye care.^14,15^

We hypothesized that a low-cost, user-friendly, clinical-grade imaging and telemedicine platform could enable non-specialist community health workers (CHWs) to capture high-quality anterior segment images, provide screening in a manner that is acceptable to patients, and achieve diagnostic accuracy comparable to in-person ophthalmologist exams. To test this, we developed Scout^TM^, a novel smartphone-based anterior segment imaging modality that standardizes image capture, illumination, magnification, and resolution through intraocular lens (IOL)-enabled optical system. We also developed a telemedicine platform including a smartphone app for CHWs to upload screening images and data to the cloud and a web dashboard for ophthalmologists to remotely perform image and data review, diagnosis, and referral. In collaboration with Johns Hopkins University and AEH Pondicherry, we validated this approach in a 1,093-patient diagnostic concordance study performed at five eye camp locations in South India, comparing diagnosis and referral agreement between CHW-led image capture with remote ophthalmologist (RO) review to conventional in-person eye camp ophthalmologist (ECO) examination.

## 2. MATERIALS AND METHODS

### 2.1 Imaging system fabrication and testing

We developed a low-cost, smartphone-based anterior segment imaging system (Scout^TM^) comprised of an IOL, light-emitting diode (LED) illumination, a custom silicone scope, and a smartphone attachment paired with a Samsung Galaxy M21 smartphone (48 MP camera, 4 GB RAM, 64 GB ROM). The 10-diopter Aurovue EV IOL (Aurolab, Madurai, India) was selected after comparative optical testing of 8 D, 10 D, and 12 D lenses using depth-of-field and resolution targets. Illumination was optimized by testing round, flat, and strip white LEDs, resulting in a configuration with two 5 mm, 3 V cool-white LEDs (Chanzon, Guangdong, China) positioned 40 mm apart and angled 45° towards the pupil to achieve uniform diffuse illumination. The silicone scope was molded using 3D-printed tooling (Ultimaker 3, Ultimaker, New York, NY) and medical-grade silicone (Dragon Skin 10, Smooth-On, Macungie, PA), and fits comfortably against the patient’s orbital rim to block ambient light and maintain a fixed working distance between the eye and the IOL. The LED lights are powered using the smartphone’s on-the-go (OTG) connector, allowing fully portable field operation.

The final device configuration was selected based on optical performance, ease of use, and qualitative feedback from ophthalmologists reviewing images captured with each prototype. Images were graded as optimal, acceptable, or poor according to focus, centration, illumination, and visibility of key anterior segment anatomic structures including the eye’s lens status. Full device specifications, optical testing procedures, and light source comparisons are described in **Supplementary Methods Section 1**.

### 2.2 Smartphone application software design, data handling, and load testing

Image capture and data collection were performed using a mobile application integrated with a cloud-based teleophthalmology platform (InSightful) adapted from an open-source telemedicine software (Intelehealth). The Android app was compatible with version 5.0 and above and was available in English and Tamil languages. The app enables CHWs to collect data on patient demographics, ocular and medical history, and visual acuity (VA) for each eye and capture anterior segment photographs of each eye for RO review. Data is transmitted via a RESTful API to a cloud-based MySQL database structured on the OpenMRS schema. Images are stored in encrypted blob storage. When network connectivity is not available, data is temporarily stored in a SQLite database on the smartphone and uploaded when connectivity is restored. ROs accessed a secure web portal displaying patient data and images. For each eye, the web app required ROs to record image quality (poor, acceptable, or optimal), lens status (mature cataract, immature cataract, clear crystalline lens, pseudophakia, aphakia, or corneal opacity preventing lens visualization), optional additional anterior segment diagnoses (such as refractive error, pterygium, active corneal infection, or inactive corneal scar), and referral details (referral decision, location, and urgency within or beyond 14 days). Platform scalability in field conditions was evaluated with Apache JMeter (version 5.6) by simulating 200-500 concurrent users uploading patient data and images. Additional details regarding software architecture, data handling protocols, and load testing methods are described in **Supplementary Methods Section 2**. The mobile and web app displays are shown in **Supplementary Figure 1**.

### 2.3 CHW recruitment and training

Twenty-nine CHWs were recruited from local paramedical programs and underwent a 3-hour structured training and certification including an introduction to ophthalmology and training on use of the imaging hardware and software. Detailed CHW recruitment and certification procedures are described in **Supplementary Methods Section 3**.

### 2.4 Usability and acceptability surveys

Usability of the platform was assessed via surveys of 10 CHWs, 40 patients, and 6 ROs adapted from validated instruments.^16^ Survey questionnaires and administration details are available in **Supplementary Methods Section 4**.

### 2.5 Diagnostic validation study

We conducted a cross-sectional diagnostic validation study at community eye camps organized by AEH across rural villages in the Pondicherry region of South India (August 2022 to June 2023). We compared diagnoses made by three ROs reviewing smartphone images (VS, RK, KB) with those made by experienced ECOs performing conventional penlight examinations. The study was approved by the AEH and Johns Hopkins IRBs and adhered to the Declaration of Helsinki. Participants aged ≥16 years who provided informed consent were included. At eye camps, CHWs collected demographic and clinical information, measured visual acuity (VA), and captured anterior eye images using Scout^TM^. Images and data were uploaded for asynchronous RO review via the web platform. ECOs independently examined each patient in-person using standard penlight examination and assigned diagnoses. ROs and ECOs used identical diagnostic categories. ROs were masked to each other’s and ECO diagnoses. Discrepancies were adjudicated via a modified Delphi process, with a senior ophthalmologist as final arbiter. The complete adjudication workflow is outlined in **Supplementary Methods Section 5**.

The primary outcome was diagnostic concordance between RO diagnoses and ECO in-person diagnoses. Secondary outcomes included image quality ratings, referral decisions, and usability/acceptability. We planned to enroll 1,000 patients (2,000 eyes), which provided sufficient power to detect concordance between Scout^TM^-based and conventional diagnoses. Assumptions, prevalence estimates, and simulation methods used for power calculations are described in **Supplementary Methods Section 5**.

Statistical analyses were performed at the eye level, accounting for clustering by patient and CHW. We calculated Cohen’s κ, percent agreement, sensitivity, specificity, PPV, and NPV. Bootstrap resampling was used to generate confidence intervals. Subgroup analyses were conducted by gender and image quality, with adjustment for multiple comparisons using the Benjamini-Hochberg procedure.^17^ Full statistical analysis specifications, including bootstrap resampling, clustering, and multiple testing adjustments, are provided in **Supplementary Methods Section 5**.

## 3. RESULTS

### 3.1 Scout^TM^ imaging system development and load testing

We evaluated different lenses, light sources, and lighting configurations incorporated to achieve standardized, diagnostic-quality visualization of the anterior segment via diffuse illumination. Images of 40 eyes were captured using either an AcrySof^®^ 8 D, Aurovue EV 10 D, or Aurovue EV 12 D IOL. The 8 D AcrySof^®^ IOL has a focal length of 32.3 mm, the greatest depth of field (27 mm), the lowest magnification (2.6x), and the lowest maximum resolution (22.7 line pairs/mm) among the three lenses. It required the fewest attempts to capture an optimal image (1.2 ± 0.5 attempts) with 89.6% of images in focus. The 12D Aurovue EV IOL, which has a shorter focal length of 19 mm, the smallest depth of field (9 mm), the highest magnification (4.9x), and the highest maximum resolution (40 line pairs/mm), required the most attempts (1.9 ± 1.3 attempts) with only 55.7% of images in focus. The 10D Aurovue EV IOL has an intermediate focal length of 21.5 mm, a depth of field of 12.7 mm, a magnification of 4.1x, and a maximum resolution of 34 line pairs/mm. It required 1.3 ± 0.7 attempts to achieve a focused image, with 74.3% of images in focus. The 8D lens required significantly fewer attempts to capture an image in comparison to the 12D lens (p<0.05), but there was no significant difference in comparison to the 10D lens (p=0.3). However, the 8D lens lacked sufficient magnification and resolution for diagnostic evaluation (**Figure 1A**). The lens could not be clearly visualized and subtle corneal changes, such as stromal haze or early pterygium, were indistinct or undetectable. In contrast, both the 10D and 12D IOLs provided adequate magnification and clarity to visualize key diagnostic features, including lens opacity type and location, corneal clarity, and the presence or extent of pterygium. Thus, the Aurovue EV 10D lens was selected, as it offered an optimal balance between ease of use, capture of focused images, and clinical utility for diagnosis and referral.

**Figure 1.**
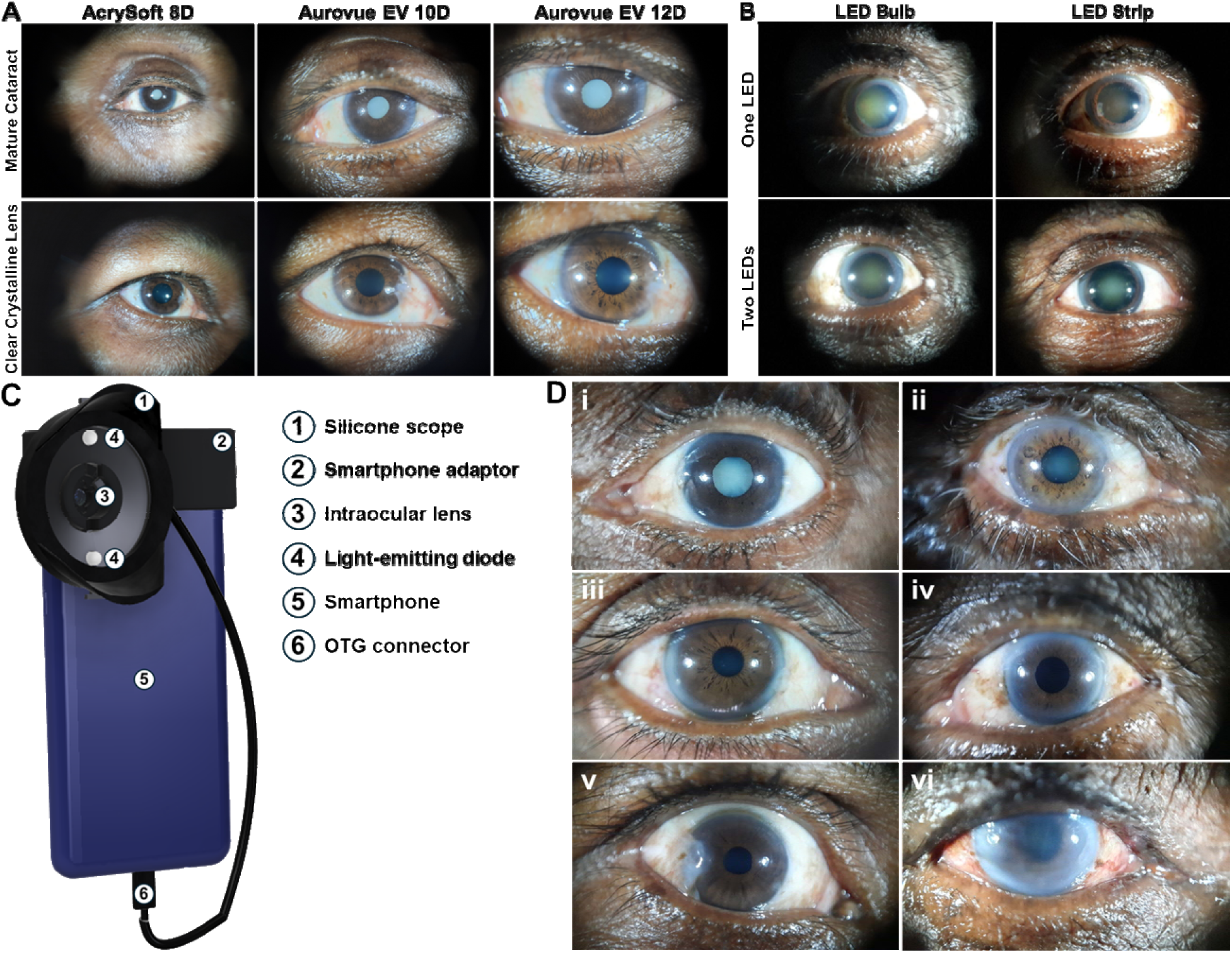
Design and validation of novel anterior segment eye imaging system. **(A)** Evaluation of IOL type and optical power in eyes with an opaque lens (mature cataract) or clear crystalline lens. **(B)** Assessment of LED number and form factor in eyes with a mature cataract. **(C)** Design and key elements of the Scout imaging system. **(D)** Representative images of anterior segment pathology captured with Scout: (i) mature cataract, (ii) immature cataract, (iii) clear crystalline lens, (iv) pseudophakia, (v) pterygium, (vi) inactive corneal opacity.

We also assessed the impact of the number and type of LEDs on visualization of the anterior segment. A single LED only partially illuminated the eye, whereas two parallel LEDs surrounding the pupil provided even illumination. The dual LED setup was preferred by ophthalmologists (53% optimal, 43% acceptable, 4% poor) in comparison to the single LED (0% optimal, 41% acceptable, 59% poor). Round LED bulbs provided uneven illumination, creating two circles of light that did not allow for appropriate visualization of the central eye area. LED strips caused large reflection artifacts that obscured important anterior eye anatomy (15% optimal, 23% acceptable, 62% poor). In contrast, flat LED bulbs offered a wider viewing angle, resulting in more even illumination and better coverage of the central eye area (**Figure 1B**). The majority of images were suitable for evaluating lens status (23% optimal, 54% acceptable, 23% poor). Thus, a dual flat LED bulb setup was selected. The resulting smartphone imaging device, Scout^TM^ (**Figure 1C**), simplifies anterior segment eye imaging by blocking ambient light, centering the eye, standardizing focal length, and optimizing lighting and magnification. Scout^TM^ enables capture of diagnostic-quality images of anterior segment pathology (**Figure 1D**), including (i) mature cataract, (ii) immature cataract, (iii) clear crystalline lens, (iv) pseudophakia, (v) pterygium, and (vi) corneal opacity, enabling screening in a range of settings.

Load testing of the mobile application demonstrated stable performance under simulated conditions of hundreds of concurrent users uploading both image and text data. These results suggested the platform could support wider deployment in community screening programs. Expanded descriptions of network load testing are presented in the **Supplementary Results Section 1**, and screenshots of the InSightful mobile phone and web app display are available in **Supplementary Figure 1**.

### 3.2 CHW performance

Throughout eye camp screenings, CHWs with limited training consistently captured high-quality images with the diffuse illumination imaging system. Across the 2,108 images captured at the eye camps, 90.7% were graded suitable for diagnosis by ROs, including 40% of images graded as “optimal” quality and 50.7% of images graded as “acceptable” quality. CHWs’ mean screening time was 8.1 ± 5.2 minutes per patient (4 minutes per eye) across all eye camps, but decreased to 4.4 ± 4.5 minutes per patient (less than 2.5 minutes per eye) in the last 4 eye camps. Details on CHW performance are presented in the **Supplementary Results Section 2**.

### 3.3 Usability and acceptability of smartphone-based screening

Surveys of CHWs, ROs, and patients demonstrated high usability and acceptability of the smartphone imaging system. Item-level responses and net promoter scores are provided in the **Supplementary Table 1** and **Supplementary Results Section 3**. CHWs were surveyed to assess the usability and acceptability of the imaging system and mobile app using a Likert scale from 1 (strongly disagree) to 5 (strongly agree) (**Table 1**). CHWs strongly agreed with the overall utility of the system to improve access to care (4.9 ± 0.3), the ease of learning (5.0) and simplicity (5.0) of the system, reliability (4.7 ± 0.5), and their confidence (4.9 ± 0.3) and satisfaction (4.7 ± 0.5) with their ability to use the system to screen patients. CHWs agreed that it was easy to take a high-quality image of the eye (4.1 ± 0.3) and collect and record information into the app (4.2 ± 0.4). We also surveyed for willingness to recommend the system to other CHWs. 80% of the CHWs were promoters and 20% were passive, resulting in an exceptional net promoter score of 80. Overall, CHWs responded with high acceptability of the screening system and its usability.

**Table 1.** Demographic and clinical characteristics of diagnostic validation study population.

| Baseline Characteristics | N |
| --- | --- |
| Participants | 1093 |
| Eyes | 2120 |
| Participant age, median (IQR) | 60 (54 – 67) |
| Participant sex, N (%) |  |
| Male | 455 (41.6) |
| Female | 638 (58.4) |
| Snellen visual acuity, meters <sup>a</sup> |  |
| 6/6 to <6/9 | 153 (7.2) |
| 6/9 to <6/24 | 646 (30.5) |
| 6/24 to <6/60 | 466 (22.0) |
| 6/60 to Hand Movements | 787 (37.1) |
| Light Perception or No Light Perception | 34 (1.6) |
| Image Quality, N (%) <sup>b</sup> |  |
| Poor | 187 (8.8) |
| Acceptable | 1083 (51.1) |
| Optimal | 846 (39.9) |
| Patient Complaints, N (%) |  |
| Blurry vision | 1787 (84.3) |
| Eye pain/irritation | 97 (4.6) |
| Watery eyes | 43 (2.0) |
| Headache | 31 (1.5) |
| Redness | 11 (0.5) |
| Light sensitivity | 2 (0.1) |

<sup>a</sup> N=34 eyes had missing Snellen visual acuity measurements. Denominator is total number of eyes. N=2 eyes had visual acuity <6/6 meters which was recategorized as 6/6.
<sup>b</sup> Denominator is total number of eyes that underwent imaging with smartphone hardware and had a quality grade assigned by a remote ophthalmologist reviewer. N=4 eyes had missing image quality grade.

ROs were surveyed on the usability of the telemedicine app and the diagnostic utility images captured with Scout^TM^. ROs strongly agreed with the simplicity of understanding patient data (4.7 ± 0.5), navigating the app (4.5 ± 0.5), inputting diagnoses (4.8 ± 0.4), the image quality being suitable for referral decisions (4.7 ± 0.5), and their confidence in the accuracy of referral made through Scout^TM^-based screening compared to penlight exam (4.3 ± 0.8). 66% of the ROs were promoters and 33% were passive, resulting in an excellent net promoter score of 66.

We conducted an acceptability survey of 40 randomly selected eye camp patients who received CHW-led smartphone-based screening (**Table 1**). Many patients traveled long distances to reach the eye camps (mean time: 40.1 ± 24 minutes; mean distance: 9.7 ± 6.4 km). Patients agreed that they were comfortable communicating their complaints to the CHW (3.9 ± 0.7), that CHWs took comprehensive notes (4.0 ± 0.8), and that it was easy to understand and follow the CHW’s directions (4.0 ± 0.6).

### 3.4 Diagnostic validation study

#### Study population

The study recruited a total of 1,202 patients (N=2,404 eyes) from 19 eye camp screenings. We analyzed N=2,120 eyes from N=1,093 patients for diagnostic concordance. **Table 1** describes the eye camp patients’ demographic characteristics and VA. The median age was 60 years (interquartile range [IQR] 54 to 67 years) and 58% of eye camp patients female. Over 60% of patients screened had distance Snellen VA of 6/24 or worse, and nearly 40% had severe visual impairment per World Health Organization (WHO) criteria. 29 CHW volunteers (67.7% female; mean age 18.3 years) were recruited from paramedical schools near camp sites. 70% of CHWs owned a smartphone.

#### Diagnosis and referral concordance by disease

**Table 2** lists the proportion of eyes with each anterior segment diagnosis as well as measures of diagnostic concordance for each diagnosis comparing in-person ECO examination (gold standard) versus CHW-led smartphone-based screening paired with RO review (new test). Lens diagnosis categories included mature cataract (7%), immature cataract (53% of eyes), clear crystalline lens (16%), pseudophakia (23%), and aphakia (N=5, 0.2%). Over 60% of eyes had any form of clinically significant cataract, whether immature or mature. Non-lens diagnoses included pterygium (6%), refractive error/presbyopia (9.5%), and inactive corneal opacity (N=14, 0.7%), and active corneal infection (N=1, 0%). Smartphone screening with RO review showed substantial agreement with ECO diagnosis for any cataract (κ 0.77, PA 88.5%), immature cataract (κ=0.69, PA 84.5%), mature cataract (κ=0.67, PA 95.6%), and clear crystalline lens (κ=0.63, PA 88.9%) and near perfect agreement for pseudophakia (κ=0.92, PA 97.2%). Percent agreement between ROs and ECOs was 84.5% for immature cataract and 87-99% for all other diagnostic categories. Compared to in-person ECO exam, the diagnostic specificity of smartphone screening with RO review was 89.1% for clear crystalline lens and 91.3-99.4% for all other diagnostic categories. RO sensitivity was 71.8% for mature cataract, 77.5% for immature cataract, and 83.8-95.8% for other lens-related diagnoses. However, measures of diagnostic concordance and sensitivity were lower for non-lens diagnoses including pterygium (κ=0.47, sensitivity 47.6%), refractive error/presbyopia (κ=0.34, sensitivity 47.5%), and inactive corneal opacity (κ=0.36, sensitivity 42.9%). Diagnostic concordance was not evaluated for aphakia (N=5), inactive corneal opacity (N=14), or active corneal infection (N=1) due to small sample size.

**Table 2.**
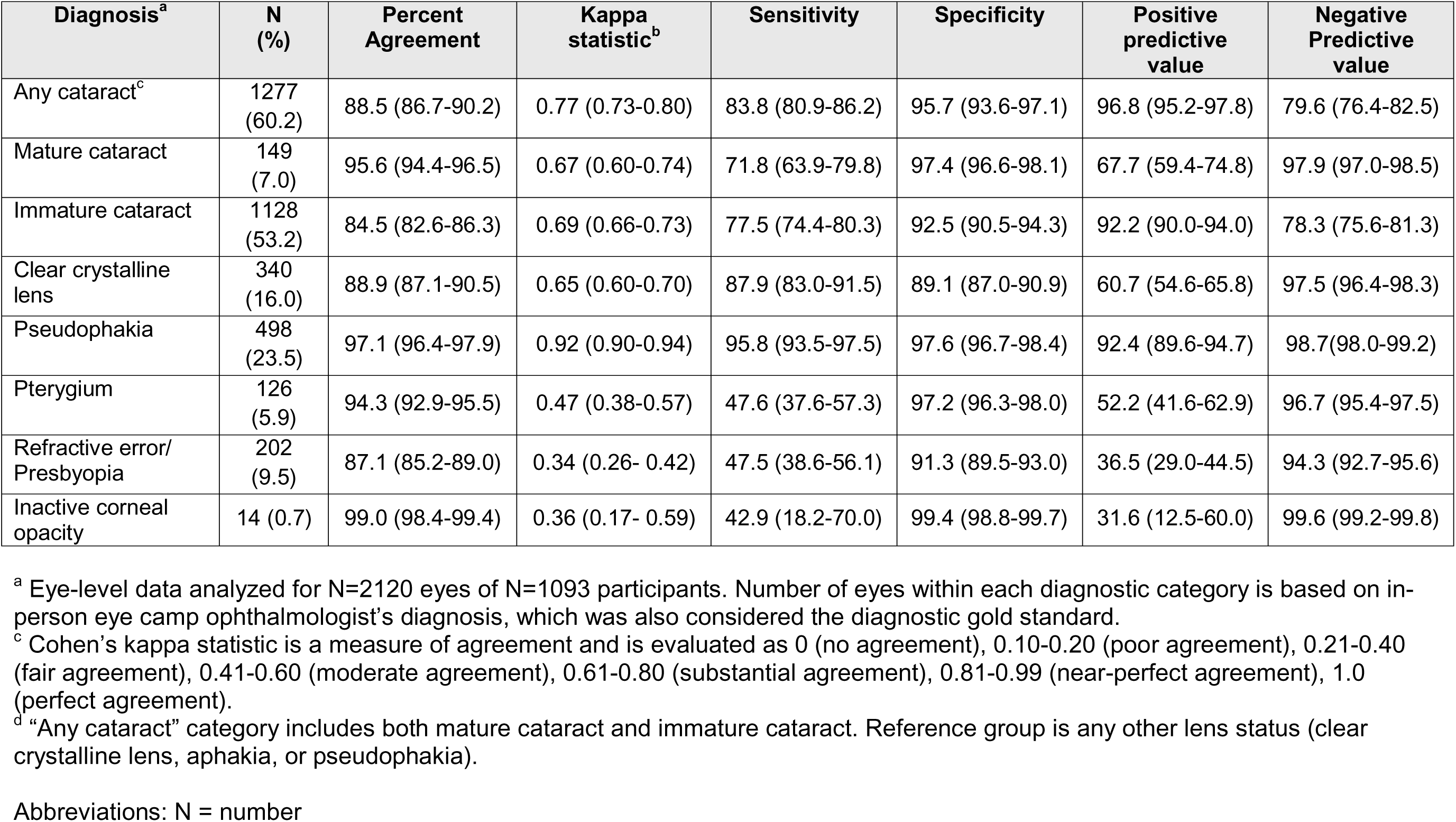
Measures of diagnostic agreement comparing remote ophthalmologist evaluation of smartphone-based screening data versus in-person eye camp ophthalmologist examination^a^.

| Diagnosis<sup>a</sup> | N (%) | Percent Agreement | Kappa statistic<sup>b</sup> | Sensitivity | Specificity | Positive predictive value | Negative Predictive value |
| --- | --- | --- | --- | --- | --- | --- | --- |
| Any cataract<sup>c</sup> | 1277 (60.2) | 88.5 (86.7-90.2) | 0.77 (0.73-0.80) | 83.8 (80.9-86.2) | 95.7 (93.6-97.1) | 96.8 (95.2-97.8) | 79.6 (76.4-82.5) |
| Mature cataract | 149 (7.0) | 95.6 (94.4-96.5) | 0.67 (0.60-0.74) | 71.8 (63.9-79.8) | 97.4 (96.6-98.1) | 67.7 (59.4-74.8) | 97.9 (97.0-98.5) |
| Immature cataract | 1128 (53.2) | 84.5 (82.6-86.3) | 0.69 (0.66-0.73) | 77.5 (74.4-80.3) | 92.5 (90.5-94.3) | 92.2 (90.0-94.0) | 78.3 (75.6-81.3) |
| Clear crystalline lens | 340 (16.0) | 88.9 (87.1-90.5) | 0.65 (0.60-0.70) | 87.9 (83.0-91.5) | 89.1 (87.0-90.9) | 60.7 (54.6-65.8) | 97.5 (96.4-98.3) |
| Pseudophakia | 498 (23.5) | 97.1 (96.4-97.9) | 0.92 (0.90-0.94) | 95.8 (93.5-97.5) | 97.6 (96.7-98.4) | 92.4 (89.6-94.7) | 98.7(98.0-99.2) |
| Pterygium | 126 (5.9) | 94.3 (92.9-95.5) | 0.47 (0.38-0.57) | 47.6 (37.6-57.3) | 97.2 (96.3-98.0) | 52.2 (41.6-62.9) | 96.7 (95.4-97.5) |
| Refractive error/ Presbyopia | 202 (9.5) | 87.1 (85.2-89.0) | 0.34 (0.26- 0.42) | 47.5 (38.6-56.1) | 91.3 (89.5-93.0) | 36.5 (29.0-44.5) | 94.3 (92.7-95.6) |
| Inactive corneal opacity | 14 (0.7) | 99.0 (98.4-99.4) | 0.36 (0.17- 0.59) | 42.9 (18.2-70.0) | 99.4 (98.8-99.7) | 31.6 (12.5-60.0) | 99.6 (99.2-99.8) |

<sup>a</sup> Eye-level data analyzed for N=2120 eyes of N=1093 participants. Number of eyes within each diagnostic category is based on in-person eye camp ophthalmologist's diagnosis, which was also considered the diagnostic gold standard.
<sup>c</sup> Cohen's kappa statistic is a measure of agreement and is evaluated as 0 (no agreement), 0.10-0.20 (poor agreement), 0.21-0.40 (fair agreement), 0.41-0.60 (moderate agreement), 0.61-0.80 (substantial agreement), 0.81-0.99 (near-perfect agreement), 1.0 (perfect agreement).
<sup>d</sup> "Any cataract" category includes both mature cataract and immature cataract. Reference group is any other lens status (clear crystalline lens, aphakia, or pseudophakia).
Abbreviations: N = number

ECOs and ROs showed strong concordance when deciding which patients required referral from the eye camp to the urban eye hospital for further care, with 96.2% agreement on patient-level referral decisions across all diagnostic categories (**Table 3**). For specific diagnoses, percent agreement on referral ranged from 91.8% for clear crystalline lens to 100% for inactive corneal opacity. Assuming ECOs to be the gold standard for referral decisions, ROs reviewing smartphone images had sensitivity of 98.6% (95% CI 97.7–99.2%) and positive predictive value of 97.4% (95% CI 96.2–98.2%) for identifying eyes that needed referral.

**Table 3:**
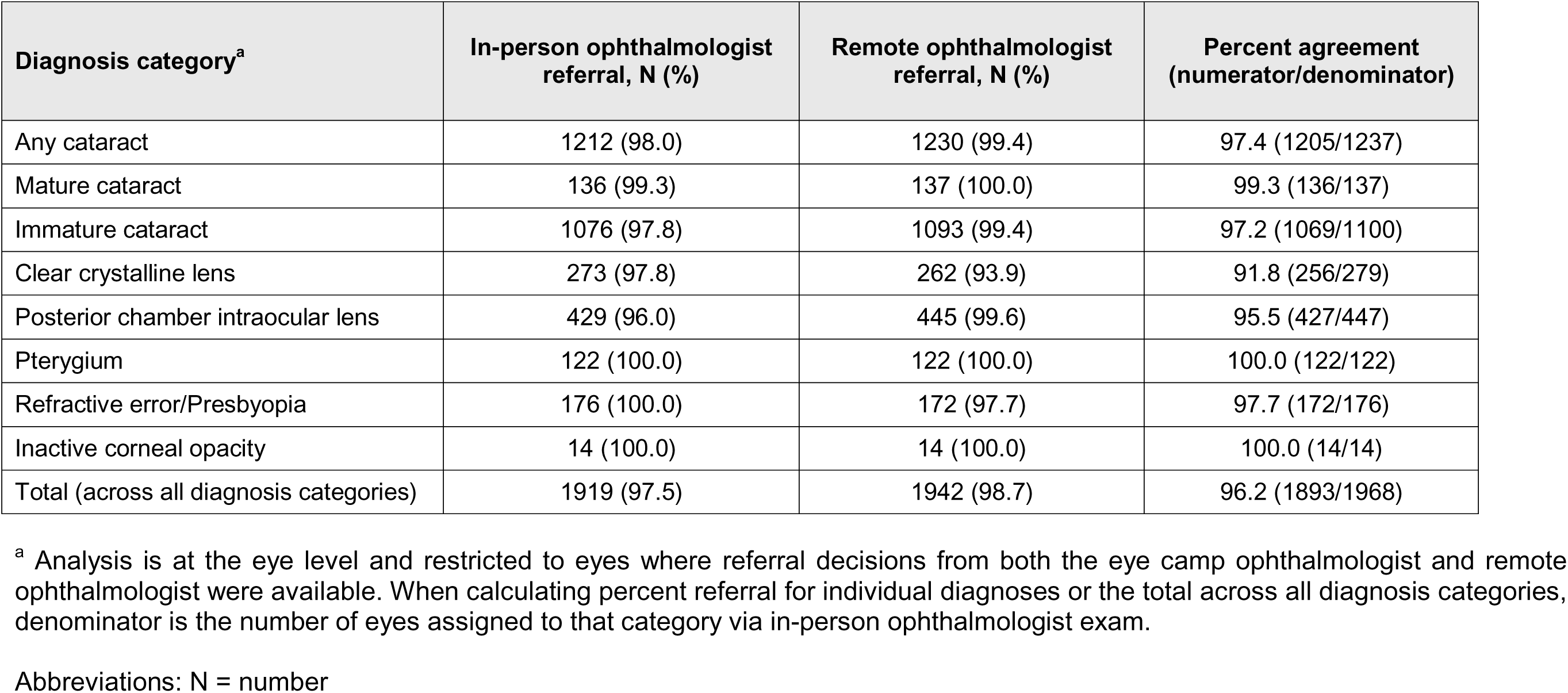
Agreement on referral decision comparing remote ophthalmologist evaluation of smartphone-based screening data versus in-person eye camp ophthalmologist examination, by diagnosis category.

| Diagnosis category<sup>a</sup> | In-person ophthalmologist referral, N (%) | Remote ophthalmologist referral, N (%) | Percent agreement (numerator/denominator) |
| --- | --- | --- | --- |
| Any cataract | 1212 (98.0) | 1230 (99.4) | 97.4 (1205/1237) |
| Mature cataract | 136 (99.3) | 137 (100.0) | 99.3 (136/137) |
| Immature cataract | 1076 (97.8) | 1093 (99.4) | 97.2 (1069/1100) |
| Clear crystalline lens | 273 (97.8) | 262 (93.9) | 91.8 (256/279) |
| Posterior chamber intraocular lens | 429 (96.0) | 445 (99.6) | 95.5 (427/447) |
| Pterygium | 122 (100.0) | 122 (100.0) | 100.0 (122/122) |
| Refractive error/Presbyopia | 176 (100.0) | 172 (97.7) | 97.7 (172/176) |
| Inactive corneal opacity | 14 (100.0) | 14 (100.0) | 100.0 (14/14) |
| Total (across all diagnosis categories) | 1919 (97.5) | 1942 (98.7) | 96.2 (1893/1968) |

<sup>a</sup> Analysis is at the eye level and restricted to eyes where referral decisions from both the eye camp ophthalmologist and remote ophthalmologist were available. When calculating percent referral for individual diagnoses or the total across all diagnosis categories, denominator is the number of eyes assigned to that category via in-person ophthalmologist exam.
Abbreviations: N = number

#### Image quality and diagnostic concordance

Diagnostic reliability improved with image quality (**Table 4**). For cataract detection, κ increased from 0.70 with poor images to 0.85 with optimal images. Odds of inter-rater concordance were more than two-fold higher when using optimal images compared with poor-quality images. Similar trends were observed for diagnosis of immature cataract and clear crystalline lens. Detailed analyses of the association of image quality with diagnostic concordance within disease classes are provided in **Supplementary Table 2** and **Supplementary Results Section 4**.

**Table 4.**
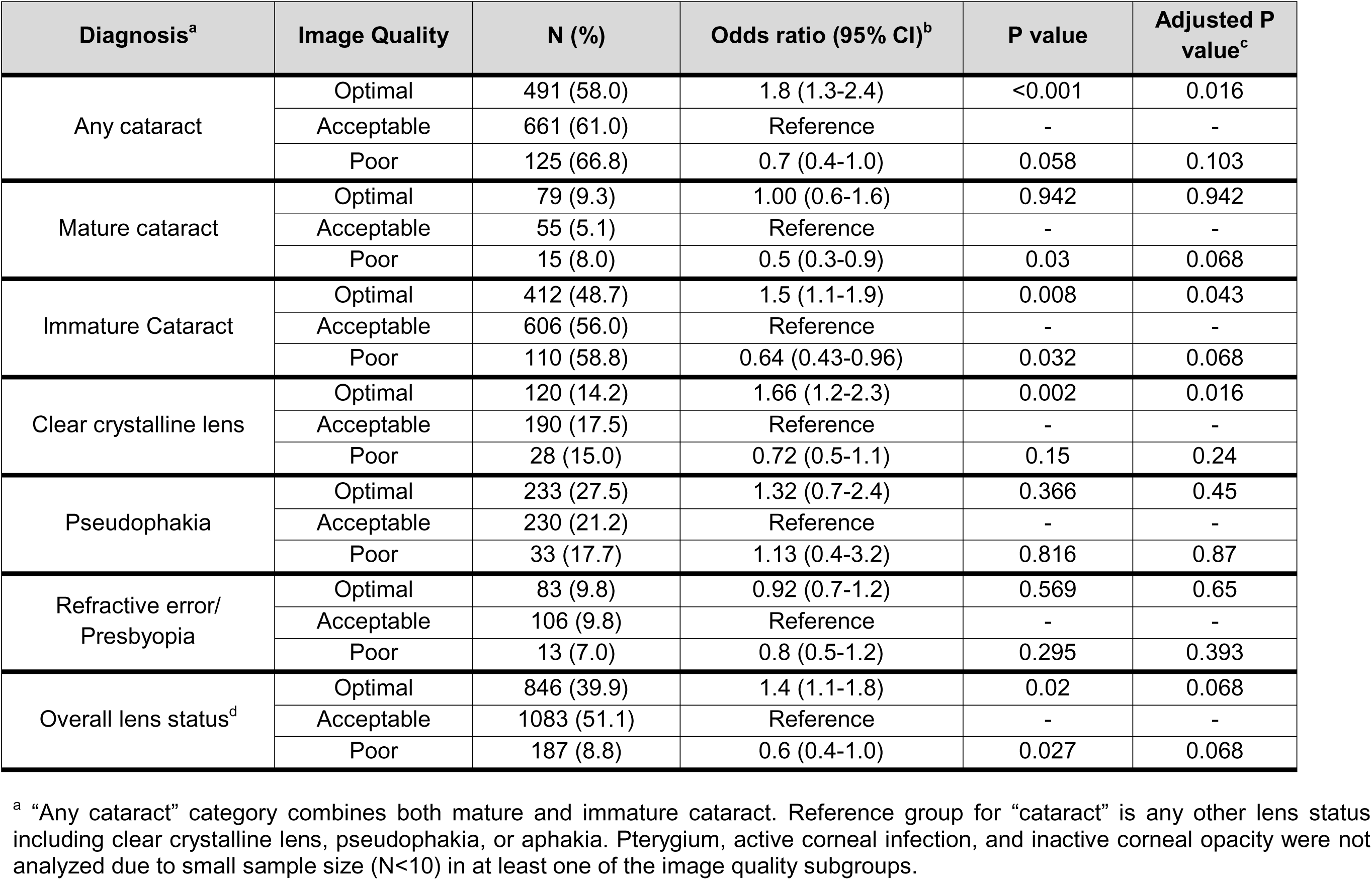
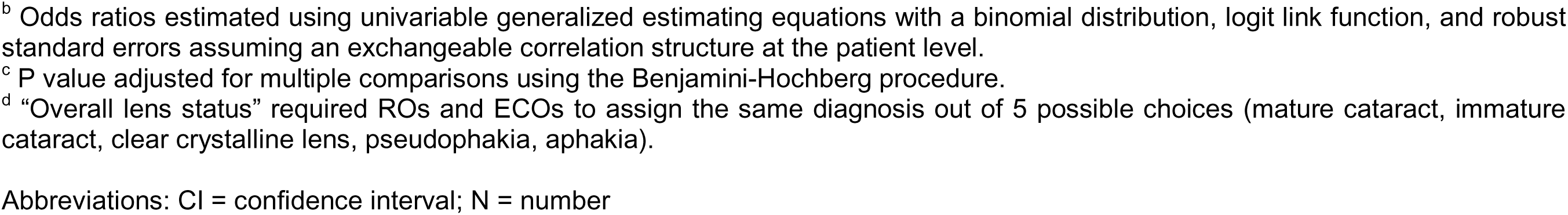
Probability of diagnostic agreement between in-person ophthalmologist examination and remote ophthalmologist evaluation of smartphone-based screening data, by image quality.

| Diagnosis<sup>a</sup> | Image Quality | N (%) | Odds ratio (95% CI)<sup>b</sup> | P value | Adjusted P value<sup>c</sup> |
| --- | --- | --- | --- | --- | --- |
| Any cataract | Optimal | 491 (58.0) | 1.8 (1.3-2.4) | <0.001 | 0.016 |
|  | Acceptable | 661 (61.0) | Reference | - | - |
|  | Poor | 125 (66.8) | 0.7 (0.4-1.0) | 0.058 | 0.103 |
| Mature cataract | Optimal | 79 (9.3) | 1.00 (0.6-1.6) | 0.942 | 0.942 |
|  | Acceptable | 55 (5.1) | Reference | - | - |
|  | Poor | 15 (8.0) | 0.5 (0.3-0.9) | 0.03 | 0.068 |
| Immature Cataract | Optimal | 412 (48.7) | 1.5 (1.1-1.9) | 0.008 | 0.043 |
|  | Acceptable | 606 (56.0) | Reference | - | - |
|  | Poor | 110 (58.8) | 0.64 (0.43-0.96) | 0.032 | 0.068 |
| Clear crystalline lens | Optimal | 120 (14.2) | 1.66 (1.2-2.3) | 0.002 | 0.016 |
|  | Acceptable | 190 (17.5) | Reference | - | - |
|  | Poor | 28 (15.0) | 0.72 (0.5-1.1) | 0.15 | 0.24 |
| Pseudophakia | Optimal | 233 (27.5) | 1.32 (0.7-2.4) | 0.366 | 0.45 |
|  | Acceptable | 230 (21.2) | Reference | - | - |
|  | Poor | 33 (17.7) | 1.13 (0.4-3.2) | 0.816 | 0.87 |
| Refractive error/<br>Presbyopia | Optimal | 83 (9.8) | 0.92 (0.7-1.2) | 0.569 | 0.65 |
|  | Acceptable | 106 (9.8) | Reference | - | - |
|  | Poor | 13 (7.0) | 0.8 (0.5-1.2) | 0.295 | 0.393 |
| Overall lens status<sup>d</sup> | Optimal | 846 (39.9) | 1.4 (1.1-1.8) | 0.02 | 0.068 |
|  | Acceptable | 1083 (51.1) | Reference | - | - |
|  | Poor | 187 (8.8) | 0.6 (0.4-1.0) | 0.027 | 0.068 |

<sup>a</sup> “Any cataract” category combines both mature and immature cataract. Reference group for “cataract” is any other lens status including clear crystalline lens, pseudophakia, or aphakia. Pterygium, active corneal infection, and inactive corneal opacity were not analyzed due to small sample size (N<10) in at least one of the image quality subgroups.
<sup>b</sup> Odds ratios estimated using univariable generalized estimating equations with a binomial distribution, logit link function, and robust standard errors assuming an exchangeable correlation structure at the patient level.
<sup>c</sup> P value adjusted for multiple comparisons using the Benjamini-Hochberg procedure.
<sup>d</sup> “Overall lens status” required ROs and ECOs to assign the same diagnosis out of 5 possible choices (mature cataract, immature cataract, clear crystalline lens, pseudophakia, aphakia).
Abbreviations: CI = confidence interval; N = number

#### Subgroup analyses

Subgroup analyses of diagnostic concordance stratified by sex are reported in the **Supplementary Table 3**. No clinically meaningful differences in diagnostic concordance were observed comparing females to males. Within each diagnostic category, difference in percent agreement between ROs and ECOs was less than 4% when comparing females versus males. RO and ECO diagnostic concordance was slightly higher for females than males for all lens-related diagnoses (p<0.001) and lower for pterygium and refractive error. Subgroup analyses stratified by age and eye camp location also did not show substantial differences across strata.

## 4. DISCUSSION

### 4.1 Smartphone screening is comparable to in-person ophthalmologist exams

We found that high-resolution diffuse illumination images captured by CHWs using the Scout^TM^ anterior segment imaging system enabled ROs to diagnose cataract and lens status with accuracy comparable to in-person eye camp ophthalmologist exams. When determining need for referral, ROs and ECOs agreed in 96% of cases. CHWs with minimal ophthalmic training rapidly achieved proficiency, captured over 90% diagnostic-quality images and completing screening in under 2.5 minutes per eye. Patients, CHWs, and ophthalmologists all reported high usability and acceptability for the system. These findings demonstrate that Scout^TM^-based anterior segment imaging and remote diagnosis has potential to effectively decentralize screening for cataract and other anterior segment disorders in low-resource environments, particularly in locations with limited access to trained eye specialists.

Prior studies have shown that remote photographic diagnosis of anterior eye diseases can achieve diagnostic performance approaching in-person ophthalmic exams.^18–20^ However, these prior studies did not perform mobile smartphone photography at the point of care, relied on trained eye care personnel rather than minimally trained CHWs to perform imaging,^5,18,20–22^ did not incorporate mobile phone uploading or RO review,^5,18–20,23,24^ did not stress test data transfer in low-bandwidth environments,^5,18–20,23,24^ and/or did not directly compare in-person versus remote ophthalmologist diagnosis.^5,22,23,25^ Perhaps most important, most prior studies have typically used eye images obtained under ideal testing settings with controlled illumination, stable patient positioning, and/or images obtained by trained eye care providers, all of which limit generalizability of results to challenging real-world LMIC settings.^5,18–20,23,24^ Our approach differs by enabling point-of-care smartphone imaging by CHWs using a low-cost, Android-compatible device optimized for field use and connectivity constraints common in LMICs. In contrast to prior work that used expensive optics or was conducted in controlled clinical environments, we designed Scout^TM^ with an inexpensive optical system and compatibility with low-cost Android phones, demonstrating a rapid learning curve for novice photographers, high acceptability among all stakeholders, and validated diagnostic performance in over 1,000 patients in low-bandwidth real-world eye camp conditions. Taken together, our results support the feasibility and scalability of this teleophthalmology platform for community eye screening in low-resource settings.

### 4.2 Modernizing eye screening beyond traditional eye camps

Eye camps have been the mainstay of cataract screening in LMICs since after World War II.^11^ Although eye camps have led to reductions in the proportion of cataract blindness and visual impairment, they have several limitations such as high cost, logistical complexity, dependence on highly trained eye specialists, restriction to pre-specified screening dates and locations, and difficulty reaching the most isolated or vulnerable patient populations such as women, immobile patients, or the very elderly.^3,12,13,26,27^ A study from AEH highlighted this disparity, finding that almost two-thirds of the rural population had never accessed any form of eye care service despite a high prevalence of ocular diseases.^28^ Smartphone-based teleophthalmology offers a more flexible, accessible, and sustainable model. By equipping CHWs with portable, high quality imaging tools and a secure telemedicine platform, screening can occur closer and more frequently to where patients live, with ophthalmologists remotely reviewing images and making timely diagnosis and referral decisions.

Our study demonstrates that a de-skilled anterior segment screening program is both feasible and efficient, producing diagnostic-quality images at scale while reducing reliance on trained ophthalmic personnel. This approach could augment or partially replace conventional eye camps, allowing more frequent and/or geographically distributed screening that alleviates human resource constraints and is integrated with existing public health infrastructure. This platform also enables the prospect of effective door-to-door screening in remote areas expanding access and helping reach the most vulnerable patients. Future research should evaluate the effectiveness of community-wide smartphone screenings for enabling accurate and timely diagnosis and referral, reducing screening costs, and improving clinical outcomes. Imaging-based anterior segment screening may also enable near-real-time diagnosis and referral using artificial intelligence (AI), further expanding access and availability.

### 4.3 Subjectivity of cataract grading

This study highlighted the inherent subjectivity of clinical cataract grading by ophthalmologists. Aravind ECOs grade cataract using binarized categories of “immature” or “mature” to evaluate candidacy for phacoemulsification versus manual small incision cataract surgery. In borderline cases, ophthalmologists can frequently disagree on whether a cataract should be classified as mature or immature. This became apparent during Delphi panel review of the images for which all 3 ROs did not agree on lens status diagnosis and engaged in robust discussions about classification. In particular, the “immature cataract” category encompassed a broad range of lens opacities between a clear crystalline lens and a fully mature cataract, including borderline cases that different graders could reasonably classify in either adjacent category. For immature cataract, clear crystalline lens, and the combined category of “any cataract” (combining both mature and immature cataract), the odds of RO and ECO concordance significantly increased with higher image quality, indicating that better illumination, focus, and/or resolution helped ROs accurately differentiate lens status. Among mature cataract, odds of diagnostic concordance did not significantly increase with high image quality, indicating that advanced cataracts are more readily diagnosable regardless of image quality. Future research should explore objective, image-based severity scoring or AI-assisted grading to improve the consistency and scalability of cataract assessment.

### 4.4 Limitations

This study had several limitations that will inform future validation. The diagnostic gold standard was in-person ophthalmologist (ECO) penlight examination at eye camps, often performed after dilation, whereas ROs graded images of undilated eyes captured by CHWs using Scout™ to avoid precipitating angle closure glaucoma in a population with high prevalence of narrow iridocorneal angles.^29^ This difference may have reduced RO accuracy for cataract grading, though high-resolution images may have enhanced RO detection of other anterior segment pathologies. ROs also reviewed images paired with visual acuity (VA) data without knowing whether VA was measured with or without spectacle correction, potentially affecting diagnostic performance. Future work should compare RO diagnosis to slit lamp ophthalmoscopy and consider latent class analysis to assess diagnostic performance without assuming a single gold standard. Future studies should also evaluate whether adding slit beam, blue light, or dilated eye images improves RO performance.

ECOs and ROs did not apply consistent diagnostic criteria for pterygium, refractive error/presbyopia, and corneal opacity. Because eye camps focused mainly on cataract screening, ECOs may have missed milder cases of pterygium or corneal opacity more easily detected by ROs reviewing high-quality images. Unlike ROs reviewing static data, ECOs could interact directly with patients and ask additional questions to determine clinical significance. Future studies should evaluate concordance for pterygium, corneal infection, corneal scarring, and other ocular surface diseases with ROs and in-person ophthalmologists applying the same diagnostic criteria. Findings from rural South Indian eye camps may not generalize to other contexts, but the short learning curve, rapid screening time, and strong usability and satisfaction ratings among CHWs and patients suggest that the Scout^TM^ device with the InSightful app may be adaptable to other settings.

## 5. CONCLUSIONS

Our novel smartphone-based anterior segment screening platform enables CHWs with minimal ophthalmic training to easily and efficiently capture clinical-grade anterior segment images and clinical data from patients in a community setting, which can then be transferred in low bandwidth settings to a remote provider for diagnosis and referral. Ophthalmologists remotely reviewing these high-resolution diffuse illumination images are able to diagnose cataract and lens status with comparable accuracy to an in-person eye camp ophthalmologist exam. Our de-skilled, decentralized platform demonstrates potential to significantly expand access to anterior eye screening for leading causes of blindness.

## Supporting information

Supplementary Section

## Data Availability

Data produced in the present study may be available upon reasonable request to the authors in line with institutional and local regulations.

## REFERENCES

1. Flaxman SR, Bourne RRA, Resnikoff S, et al. Global causes of blindness and distance vision impairment 1990-2020: a systematic review and meta-analysis. Lancet Glob Health. 2017;5(12):e1221–e1234. doi:10.1016/S2214-109X(17)30393-5

2. World report on vision. Geneva: World Health Organization; 2019. Licence: CC BY-NC-SA 3.0 IGO.

3. Burton MJ, Ramke J, Marques AP, et al. The Lancet Global Health Commission on Global Eye Health: vision beyond 2020. The Lancet Global Health. 2021;9(4):e489–e551. doi:10.1016/S2214-109X(20)30488-5

4. Whitcher JP, Srinivasan M, Upadhyay MP. Corneal blindness: a global perspective. Bull World Health Organ. 2001;79(3):214–221.

5. Goel R, Macri C, Bahrami B, Casson R, Chan WO. Assessing the subjective quality of smartphone anterior segment photography: a non-inferiority study. Int Ophthalmol. 2023;43(2):403–410. doi:10.1007/s10792-022-02437-9

6. Woodward MA, Niziol LM, Musch DC, Lee PP. Limitations of Portable Cameras for Detecting Anterior Segment Pathology. Invest Ophthalmol Vis Sci. 2017;58(8):3541.

7. Resnikoff S, Lansingh VC, Washburn L, et al. Estimated number of ophthalmologists worldwide (International Council of Ophthalmology update): will we meet the needs? Br J Ophthalmol. 2020;104(4):588–592. doi:10.1136/bjophthalmol-2019-314336

8. Courtright P, Mathenge W, Kello AB, Cook C, Kalua K, Lewallen S. Setting targets for human resources for eye health in sub-Saharan Africa: what evidence should be used? Hum Resour Health. 2016;14:11. doi:10.1186/s12960-016-0107-x

9. Bastawrous A, Hennig BD. The global inverse care law: a distorted map of blindness. Br J Ophthalmol. 2012;96(10):1357–1358. doi:10.1136/bjophthalmol-2012-302088

10. The State of Mobile Internet Connectivity 2024 | GSMA Intelligence. October 23, 2024. Accessed October 5, 2025. https://www.gsmaintelligence.com/research/the-state-of-mobile-internet-connectivity-2024

11. Gupta SK, Murthy GVS. Where Do Persons With Blindness Caused by Cataracts in Rural Areas of India Seek Treatment and Why? Archives of Ophthalmology. 1995;113(10):1337–1340. doi:10.1001/archopht.1995.01100100125046

12. Fletcher AE, Donoghue M, Devavaram J, et al. Low Uptake of Eye Services in Rural India: A Challenge for Programs of Blindness Prevention. Archives of Ophthalmology. 1999;117(10):1393–1399. doi:10.1001/archopht.117.10.1393

13. Finger RP, Ali M, Earnest J, Nirmalan PK. Cataract surgery in Andhra Pradesh state, India: an investigation into uptake following outreach screening camps. Ophthalmic Epidemiol. 2007;14(6):327–332. doi:10.1080/01658100701486814

14. Chong JC, Tan CHN, Chen DZ. Teleophthalmology and its evolving role in a COVID-19 pandemic: A scoping review. Ann Acad Med Singap. 2021;50(1):61–76.

15. Sharma M, Jain N, Ranganathan S, et al. Tele-ophthalmology: Need of the hour. Indian J Ophthalmol. 2020;68(7):1328–1338. doi:10.4103/ijo.IJO_1784_20

16. Parmanto B, Lewis AN, Graham KM, Bertolet MH. Development of the Telehealth Usability Questionnaire (TUQ). Int J Telerehabil. 2016;8(1):3–10. doi:10.5195/ijt.2016.6196

17. Benjamini Y, Hochberg Y. Controlling the False Discovery Rate: A Practical and Powerful Approach to Multiple Testing. Journal of the Royal Statistical Society Series B (Methodological). 1995;57(1):289–300.

18. Yazu H, Shimizu E, Okuyama S, et al. Evaluation of Nuclear Cataract with Smartphone-Attachable Slit-Lamp Device. Diagnostics (Basel). 2020;10(8):576. doi:10.3390/diagnostics10080576

19. Ghafarian S, Masoumi A, Tabatabaei SA, et al. Clinical evaluation of corneal ulcer with a portable and smartphone-attachable slit lamp device: Smart Eye Camera. Sci Rep. 2025;15(1):3099. doi:10.1038/s41598-025-87820-z

20. Duong NM, Nguyen Vu NQ, Le HT. Diagnostic Assessment of Nuclear Cataracts Using a Smartphone-Attachable Slit-Lamp Device: A Cross-Sectional Study in Vietnam. Cureus. 2024;16(11):e73783. doi:10.7759/cureus.73783

21. Collon S, Chang D, Tabin G, Hong K, Myung D, Thapa S. Utility and Feasibility of Teleophthalmology Using a Smartphone-Based Ophthalmic Camera in Screening Camps in Nepal. Asia Pac J Ophthalmol (Phila). 2020;9(1):54–58. doi:10.1097/01.APO.0000617936.16124.ba

22. Mercado C, Welling J, Oliva M, et al. Clinical Application of a Smartphone-Based Ophthalmic Camera adapter in Under-Resourced Settings in Nepal. J Mob Technol Med. 2017;6(3):34–42. doi:10.7309/jmtm.6.3.6

23. Hu S, Wu H, Luan X, et al. Portable Handheld Slit-Lamp Based on a Smartphone Camera for Cataract Screening. Journal of Ophthalmology. 2020;2020:1–6. doi:10.1155/2020/1037689

24. Triningrat AAMP, Doniho A, Jayanegara WG, et al. Reliability and Accuracy of Smart Eye Camera in Determining Grading of Nuclear Cataract. Korean J Ophthalmol. Published online February 26, 2025. doi:10.3341/kjo.2023.0131

25. Ludwig CA, Newsom MR, Jais A, Myung DJ, Murthy SI, Chang RT. Training time and quality of smartphone-based anterior segment screening in rural India. Clin Ophthalmol. 2017;11:1301–1307. doi:10.2147/OPTH.S134656

26. Aboobaker S, Courtright P. Barriers to Cataract Surgery in Africa: A Systematic Review. Middle East Afr J Ophthalmol. 2016;23(1):145–149. doi:10.4103/0974-9233.164615

27. Mailu EW, Virendrakumar B, Bechange S, Jolley E, Schmidt E. Factors associated with the uptake of cataract surgery and interventions to improve uptake in low- and middle-income countries: A systematic review. PLOS ONE. 2020;15(7):e0235699. doi:10.1371/journal.pone.0235699

28. Nirmalan PK, Katz J, Robin AL, et al. Utilisation of eye care services in rural south India: the Aravind Comprehensive Eye Survey. British Journal of Ophthalmology. 2004;88(10):1237–1241. doi:10.1136/bjo.2004.042606

29. Vijaya L, George R, Arvind H, et al. Prevalence of Angle-Closure Disease in a Rural Southern Indian Population. Arch Ophthalmol. 2006;124(3):403–409. doi:10.1001/archopht.124.3.403

