## Supplementary Section for "Development and validation of a user-friendly smartphone imaging and telemedicine platform for remote diagnosis of anterior segment eye disease"

**SUPPLEMENTARY MATERIAL**

### **SUPPLEMENTARY METHODS**

This section contains detailed descriptions of methods used for smartphone device design and testing, community health worker training, stakeholder surveys, and study design, power and sample size calculations, and statistical analyses for the diagnostic validation study.

#### **1. Imaging system fabrication and testing**

Scout^TM^ consists of a magnifying lens, light source, scope, and smartphone attachment. Scout^TM^ was used in conjunction with a Samsung Galaxy M21 smartphone (48 megapixel camera) which was selected based on cost, processing speed, camera megapixels, and availability in India. Samsung Galaxy M21 2021 Edition (SM-M215G/DS) smartphones with 64 GB ROM and 4 GB RAM were procured from Samsung India Electronics Pvt. Ltd. (New Delhi, India). Scout^TM^ is compatible with smartphones with at least 2 gigabytes RAM and a 12 megapixel camera. We evaluated three monofocal IOLs including an AcrySof^®^ SA60AT 8 diopter (D) lens (Alcon, Fort Worth, TX), an Aurovue EV 10 D lens (Aurolab, Madurai, Tamil Nadu, India), and an Aurovue EV 12 D lens. For each lens, the focal length and depth of field were measured using a DOF 5-15 Depth of Field Target (Edmund Optics, Barrington, NJ), and the resolution was measured using a 1951 USAF Resolution Target (n=3; Edmund Optics, Barrington, NJ). At AEH, a trained technician captured images of 40 eyes from 20 patients with each lens in random order. We measured the number of imaging attempts needed to capture an in-focus image, defined as >50% of the pupillary margin appearing in sharp focus. To assess magnification, the size of the eye was measured in ImageJ for each lens type (8D, 10D, and 12D) (n=20). Images were reviewed by four ophthalmologists, followed by qualitative interviews regarding their diagnostic utility.

We evaluated three different light sources including round light-emitting diode (LED) bulbs, flat LED bulbs, and LED strips in 12 eyes from 6 patients (3 male, 3 female). White LEDs with flat lenses (29 mm long lead, DC 3V) were acquired from EDGELEC (Guangdong, China). 5 mm white LEDs (clear round transparent, DC 3V 20 mA) were purchased from Chanzon (Guangdong, China). Natural white (4000K) LED strip lights were obtained from KXZM (Sichuan, China). For each lighting embodiment, an image was captured with the left LED, the right LED, or both LEDs. Resulting image quality and diagnosability were qualitatively assessed by three ophthalmologists. Images were graded as optimal, acceptable, or poor based on overall quality, including focus, centering, illumination, and the visibility of key anatomical structures. Optimal images were well-focused, centered, and evenly illuminated, with the lens status clearly visible and suitable for clinical interpretation. Acceptable images had minor issues, such as slight blur, off-centering, or uneven lighting, but lens status could still be assessed. Poor images were significantly degraded, preventing visualization of lens status, and not suitable for diagnosis.

The imaging system used in the validation study consists of a 10D Aurovue EV IOL positioned in front of the smartphone camera aperture and two white LEDs (5 mm 3V Cool White) spaced evenly 40 mm apart to the left and right of the pupil and angled at 45 degrees into the eye. The LEDs are powered by the smartphone on-the-go (OTG) connector for portability. A soft silicone scope fits against the orbital cavity to block external lighting and standardizes the 25 mm distance from the eye to the IOL. For the silicone scope, a two-part mold was 3D-printed using an Ultimaker 3 (Ultimaker, New York, NY) with black PLA PRO (PLA+) filament from eSun (Shenzhen, China) and Dragon Skin 10 Medium silicone from Smooth-On (Macungie, PA).

#### **2. Smartphone application software design, data handling, and load testing**

The InSightful platform for remote image/data review was adapted from Intelehealth (Thane, Maharashtra, India), an open-source telemedicine software. InSightful consists of an Android mobile app for CHWs to screen patients and a web app for ROs to review patient information and images to make a diagnosis. The mobile app is compatible with Android 5.0 and above; is available in English and Tamil; collects data on patient demographics, eye-related complaints, basic medical and ocular history, and visual and pinhole acuity; and facilitates ocular image capture. Data is uploaded to a cloud-based MySQL database and images are uploaded to cloud-based blob storage via a RESTful API. The MySQL database is structured using the OpenMRS data model, an open-source medical record system. When network connectivity is unavailable, data is temporarily stored in a SQLite database on the mobile device and uploaded once connectivity is restored. The web app displays a summary of each visit, including patient complaints, history, visual acuity, and anterior eye images for RO review. For each patient visit, the web app requires the RO to input image quality for each eye (optimal, acceptable, or poor), diagnosis for each eye, referral decision, referral location, and referral urgency (follow-up at eye hospital recommended within or beyond 14 days). For diagnosis of lens status, it is mandatory to select a lens status of mature cataract, immature cataract, clear crystalline lens, pseudophakia, aphakia, or corneal opacity preventing lens visualization. It is optional to input one or more additional diagnoses such as refractive error, pterygium, active corneal infection, inactive corneal scar/opacity, or suspected posterior segment pathology. **Supplementary Figure 1** shows the mobile and web app displays.

To evaluate performance and scalability under simulated field conditions, load testing using Apache JMeter (version 5.6) assessed the mobile app’s ability to upload ocular images and text-based patient data. Three scenarios were evaluated: short-term increased usage (300 concurrent users over five consecutive upload cycles), medium-term increased usage (200 concurrent users over ten consecutive upload cycles), and small spikes in usage (more than 500 concurrent users uploading simultaneously). Each simulated user session involved a full upload of patient demographics, clinical complaints, basic medical and ocular history, visual acuity measurements, and two ocular images per visit. Text data payloads averaged approximately 10 kB per upload, while image uploads averaged 2 MB. The error rate was measured for both text and image uploads.

#### **3. CHW recruitment and training**

CHW volunteers were recruited from paramedical training programs located near camp sites. All CHWs had completed high school, were fluent in Tamil, and were first-year paramedical students. CHWs were trained through a workshop consisting of three one-hour modules: (1) Introduction to eye care, ocular anatomy, common ocular diseases, and diseases requiring emergency referral; (2) Navigating the mobile app and collecting medical history and complaints; (3) Taking an image of the eye using Scout^TM^ and assessing whether the image is suitable for diagnosis. Afterwards, CHWs screened 10 patients at an eye camp and were evaluated based on four pass/fail criteria: interaction quality, image quality, safety, and emergency referral accuracy. To receive a passing mark for interaction quality, CHWs needed to clearly introduce themselves and explain the purpose of the study to the patient. For image quality, both eye images had to be graded by an ophthalmologist as sufficiently high quality to enable clinical diagnosis of the ocular condition. Image quality was graded as poor (not enabling clinical diagnosis by an RO), fair (enabling clinical diagnosis), or good (enabling confident clinical diagnosis). For safety, CHWs were required to sanitize the imaging device and their hands between each patient. For emergency referral accuracy, CHWs had to correctly identify conditions such as red eye, pus, ocular trauma, or hypermature cataract. To achieve certification, each CHW was required to achieve passing marks in all categories for at least 80% of patients screened. CHW performance was monitored throughout the study period.

#### **4. Usability and acceptability surveys**

After two eye camps, an oral survey was administered to 10 randomly selected CHWs (70% female, mean age 19.0 years). The survey was adapted from the Telehealth Usability Questionnaire (TUQ) to assess the usability and acceptance of the mobile application and eye imaging system.^1^ CHWs rated their agreement with each statement on a five-point Likert scale ((1): strongly disagree to (5): strongly agree) to gauge usefulness, ease of use, effectiveness, reliability, and satisfaction. The survey also assessed net promoter score (NPS). A separate oral survey for acceptability was administered to 40 randomly selected patients at Arani and Chengam eye camps (57.5% female, mean age 55.9). The survey assessed patient comfort communicating with CHWs and the time and distance traveled to reach the eye camp location. Patient comfort was recorded on a five-point Likert scale. Screening time was automatically recorded by the app based on the start and end timestamps of the patient encounter.

#### **5. Diagnostic validation study**

*Comparison of ECO versus RO diagnoses*

All ECOs and the three ROs performing image review (VS, RK, KB) were senior ophthalmology residents, fellows, or faculty at AEH with extensive experience performing eye camp screenings. Visual acuity was measured at eye camps using a paper Snellen chart. ECOs then performed pen light exams and assigned a diagnosis to each eye. The categories for diagnosis included mature cataract, immature cataract, clear crystalline lens, pseudophakia, aphakia, refractive error/presbyopia, pterygium, inactive corneal opacity, and active corneal opacity. Smartphone-based screening included visual acuity assessment using a paper Snellen chart, imaging performed by CHWs using Scout, and remote asynchronous image and data review and diagnosis of each eye by three independent ROs. Diagnosis categories for ROs and ECOs were identical. ROs were masked to each other’s and ECO diagnoses. If all three ROs did not agree on diagnosis for an eye, a modified Delphi panel was used to resolve disagreements. In cases where the modified Delphi panel did not reach consensus, a fourth senior ophthalmologist (NSS) masked to ROs’ diagnoses reviewed the images and rendered a decision. Data from N=170 eyes was excluded from analysis due to missing demographic/clinical information (N=114 eyes), missing images for ROs review (N=50 eyes), and non-gradable image quality (N=6 eyes). Data from an additional N=114 eyes was excluded due to missing ECO diagnosis (N=109 eyes) and RO diagnosis (N=5 eyes), respectively.

*Power and sample size calculations*

Assuming analysis at eye level and clustering of diagnoses at patient and CHW levels, an average enrollment of 50 participants (m=100 eyes) within each of 20 clusters (CHWs) was estimated. To determine eye-level prevalence of specific diagnoses, study investigators reviewed 520 eye camp diagnoses. The intra-cluster correlation coefficient (ρ) was estimated using simulations by generating 20 clusters of 50 bivariate binary data points within each cluster, assuming eye-level prevalence of diseases varying from 1% (pterygium) to 53% (cataract) and between-eye correlation ranging from 0.01 to 0.8 for a diagnosis. The design effect (DE) was then estimated as DE=1+(m-1)*ρ. Given the anticipated enrollment of 1,000 patients (2,000 eyes), the effective sample size (i.e., the sample size if all eyes were independent), calculated as 2,000/DE, ranged from 496 to 787 eyes. An effective sample size between 450 and 850 for subsequent calculation of minimum detectable concordance proportions was used. Assuming alpha=0.05, power=0.9, and a null hypothesis of 60-90% concordance between remote ophthalmologists diagnoses and eye camp diagnoses, using a two-sided test for one sample proportion, the minimum detectable concordance proportions for CHW/RO vs ECO diagnoses ranged from 0.65 to 0.94. A sample size of 1,000 patients (2,000 eyes) and 20 CHWs was estimated to provide sufficient power.

*Statistical analysis*

To assess differences in performance between Scout^TM^ lenses, a repeated measures ANOVA was conducted to compare the mean number of attempts required across the three lenses. Post-hoc pairwise comparisons were performed using Tukey’s Honestly Significant Difference (HSD) test. We assessed diagnostic concordance using Cohen’s kappa statistic (κ; measure of agreement after accounting for agreement occurring due to chance alone^2^), percent agreement (PA), sensitivity, specificity, positive predictive value (PPV), and negative predictive value (NPV). All analyses were conducted at eye level. We performed subgroup analyses by gender and image quality. Primary and subgroup analyses were not conducted when the diagnosis category appeared in 10 or fewer eyes. Confidence intervals for diagnostic accuracy were generated using the bias-corrected and accelerated (BCa) bootstrap method. We checked for correlations at patient, CHW, and eye camp level using interclass correlation coefficients (ICCs). Due to high ICC scores (any cataract 0.488; mature cataract 0.482; immature cataract 0.485; clear crystalline lens 0.995; pseudophakia 0.259; pterygium 0.971; refractive error/presbyopia 0.995; inactive corneal opacity 0.609), clustered bootstrapping was used to resample with replacement at the patient level. Each bootstrap was initialized by setting a random seed and running 1,000 iterations. To examine levels of RO-ECO concordance across different gender or image quality subgroups, we fit a univariable generalized estimating equation (GEE) model using binomial family, logit link, and robust variance estimation assuming exchangeable correlation structure at the patient level. We compared concordance estimates between subgroups using the empirical distribution of estimates obtained by running BCa bootstrap iterations. Normality was checked using the Shapiro-Francia test. An unpaired t-test was used to compare means between groups that followed a normal distribution. Non-parametric tests were used for non-normal distributions. Due to multiple comparisons, p-values were adjusted using the Benjamini-Hochberg procedure. P-values of <0.05 and confidence intervals of odds ratios that did not overlap the null value (=1) were considered statistically significant. All statistical analyses were performed using Stata 17.0 (StataCorp LLC, College Station, TX). Continuous baseline variables are presented as mean (or median, depending on distribution) and categorical variables as percentages.

### **SUPPLEMENTARY RESULTS**

#### **1. Scout^TM^ load testing**

*InSightful network load testing*

Load testing demonstrated stable performance of the InSightful platform across all simulated usage scenarios. During short-term increased usage (300 concurrent users over five upload cycles), the text and image upload functions maintained low error rates of 0.067% each. Under medium-term increased usage (200 concurrent users over ten cycles), error rates were further reduced to 0.05% for both text and image uploads. During the simulated spike scenario, involving over 500 concurrent users, the text upload error rate rose modestly to 0.1%, while the image upload error rate increased to 0.38%. Despite increased concurrency, the platform consistently achieved high upload success rates with minimal degradation in performance, indicating robust scalability of the cloud-based MySQL database and blob storage systems under variable network and load conditions likely to be experienced in low resource settings.

#### **2. CHW performance**

CHWs were able to quickly achieve proficiency with both Scout^TM^ and the InSightful platform. Twenty-two of 29 CHWs were successfully certified to participate in the eye camp diagnostic validation study by obtaining ≥80% high-quality images during the initial 4-hour training workshop. Three CHWs failed to achieve an 80% high-quality image threshold during the first eye camp, but exceeded this threshold after an additional one-hour training session. The 22 CHWs performing eye camp screenings achieved an average of 88.3 ± 7.2% high-quality images during their first eye camp screening session.

CHWs were also able to efficiently perform screenings, including collection of patient information and complaints, ocular image capture, and information input into the mobile app. The mean screening time across all visits was 8.06 minutes. Screening duration decreased over the course of the study, indicating improved efficiency among CHWs. A linear regression demonstrated a significant downward trend in average screening time by date (slope = –0.57 minutes/eye camp, intercept = 14.46 minutes, R^2^ = 0.7264), suggesting that each subsequent eye camp was associated with an average reduction in screening time of 34 seconds. The average screening time at the beginning of the study period was approximately 14.5 minutes, compared to less than 5 minutes by the end of the study period.

#### **3. Image quality and diagnostic concordance**

Image quality was graded by ROs as optimal (N=891, 40.0%) or acceptable (N=1131, 50.7%) for clinical diagnosis in 90.7% of eyes. Image quality was graded as poor in N=208 eyes (9.3%). **Supplementary Table 2** presents measures of diagnostic concordance for each diagnosis category by image quality. Except for percent agreement (P value=0.672) and specificity (P value=0.892) of pseudophakia, all diagnostic concordance parameters showed statistically significant differences among optimal, acceptable, and poor-quality images with trends toward improved diagnostic performance with higher image quality (all P value<0.001). The only exceptions were percent agreement and positive predictive value for pseudophakia, which were consistently high at 96-97% regardless of image quality grade. **Table 4** presents odds of diagnostic agreement between ECOs and ROs by image quality, using the “acceptable” image quality grade as the reference group. Compared to acceptable quality images, optimal quality images reviewed by ROs had 45-75% higher odds of being assigned the same diagnosis as ECOs for immature cataract (OR_optimal vs. acceptable_ 1.45, 95% CI 1.10–1.92), any cataract (OR_optimal vs. acceptable_ 1.75, 95% CI 1.28–2.39), and clear crystalline lens (OR_optimal vs. acceptable_ 1.66, 95% CI 1.21–2.29). Odds of diagnostic agreement between ROs and ECOs were 36-50% lower with poor image quality for mature cataract (OR_poor vs. acceptable_ 0.49, 95% CI 0.25–0.93) and immature cataract (OR_poor vs. acceptable_ 0.64, 95% CI 0.43–0.96). We also evaluated odds of ROs and ECOs assigning an eye the exact same “lens status” diagnosis out of five possible choices (mature cataract, immature cataract, clear crystalline lens, pseudophakia, or aphakia). Odds of ROs agreeing with ECOs on lens status were 36% lower when reviewing poor quality images (OR _poor vs. acceptable_ 0.64, 95% CI 0.43 – 0.01, P value 0.027, adjusted P value 0.068) and 38% higher when reviewing optimal quality images (OR_optimal vs. acceptable_ 1.38, 95% CI 1.05 – 1.80, P value 0.020, adjusted P value 0.068). Diagnostic concordance for pseudophakia and refractive error/presbyopia were not associated with image quality.

### **SUPPLEMENTARY TABLES**

#### **Supplementary Table 1. Community health worker, eye camp patient, and remote ophthalmologist survey responses**

| **Stakeholder group** | **Questions** | **Mean ± SD** |
| --- | --- | --- |
| **Community health workers (N=10)** | **Usefulness^a^** |  |
|  | This system will improve access to care for patients | 4.9 ± 0.3 |
|  | This system will save patients time compared to traveling to an optometrist or ophthalmologist | 4.5 ± 0.4 |
|  | **Ease of use^a^** |  |
|  | It was simple to use this system | 5.0 |
|  | It was easy to learn how to use the system | 5.0 |
|  | I was able to become productive quickly using the system | 4.7 ± 0.5 |
|  | Collecting and recording information was easy | 4.2 ± 0.4 |
|  | Taking a high-quality image was easy | 4.1 ± 0.3 |
|  | Measuring visual acuity was easy | 3.5 ± 0.9 |
|  | After training, I feel confident in my ability to screen patients | 4.9 ± 0.3 |
|  | **Interface quality^a^** |  |
|  | The app is simple and easy to understand | 4.6 ± 0.5 |
|  | **Reliability^a^** |  |
|  | Whenever I made a mistake, I could easily recover from it | 4.7 ± 0.5 |
|  | **Satisfaction^a^** |  |
|  | I am satisfied with my ability to screen patients with the  system | 4.7 ± 0.5 |
|  | **Net promoter score^b^** |  |
|  | How likely are you to recommend this to another  community health worker? (1-10) | 9.6 ± 0.8 |
| **Eye camp patients (N=40)** | I feel comfortable when communicating my complaints with the screener | 3.9 ± 0.7 |
|  | I felt the screener took notes of all my concerns | 4.0 ± 0.8 |
|  | It was easy to follow and understand the directions given by the screener | 4.0 ± 0.6 |
| **Remote ophthalmologists (N=6)** | It is simple to understand the data gathered at a patient’s screening visit. | 4.7 ± 0.5 |
|  | It is simple to navigate between patients using the application | 4.5 ± 0.5 |
|  | I am easily able to input notes and patient diagnosis with the AROMA application | 4.8 ± 0.4 |
|  | I could become productive quickly using the application | 4.8 ± 0.4 |
|  | The images of patients’ eyes are of adequate quality for me to make an appropriate referral | 4.7 ± 0.5 |
|  | I have the information I need to make a reliable decision on patient referral | 4.0 ± 1.1 |
|  | I feel confident in the accuracy of referrals made through the telemedicine system compared to in-person eye camp exams | 4.3 ± 0.8 |
|  | This system has the features or tools I need to make a referral decision | 4.2 ± 1.0 |
|  | How likely are you to recommend this platform to a colleague? | 9.2 ± 1.3 |

^a^ Community health workers were asked structured questions about the screening platform using the Telehealth Usability Questionnaire

^b^ Net promoter score is a measure of the proportion of community health workers that would recommend the screening platform to others. The average score on a scale of 1 to 10 was was 9.6 ± 0.8 across all CHWs surveyed. When classified into promoter or passive responses, 80% of community health workers were promoters and 20% were passive, resulting in a net promoter score of 80 for all CHWs surveyed.

Abbreviations: N = number; SD = standard deviation

#### **Supplementary Table 2. Measures of diagnostic agreement between remote ophthalmologist evaluation of smartphone-based screening data versus eye camp ophthalmologist in-person examination, by image quality^a^**

| **Diagnosis^b^** | **Image Quality^c^** | **N (%)^c^** | **Percent agreement** | **Kappa statistic^d^** | **Sensitivity** | **Specificity** | **Positive predictive value** | **Negative predictive value** |
| --- | --- | --- | --- | --- | --- | --- | --- | --- |
| Any cataract^c^ | Poor | 125 (66.8) | 79.7 (72.8-86.3) | 0.59 (0.45-0.71) | 73.6 (63.8-82.4) | 91.9 (77.3-98.4) | 94.9 (85.7-99.0) | 63.3 (51.3-74.7) |
|  | Acceptable | 661 (61.0) | 88.0 (85.7-90.1) | 0.76 (0.71-0.80) | 83.2 (79.5-86.1) | 95.5 (92.9-97.4) | 96.7 (94.5-98.0) | 78.4 (74.0-82.2) |
|  | Optimal | 491 (58.0) | 91.1 (88.3-93.3) | 0.82 (0.77-0.86) | 87.2 (83.1-90.5) | 96.6 (93.0-98.5) | 97.3 (94.3-98.7) | 84.5 (79.7-88.5) |
|  | P value^e^ |  | 0.0001 | 0.0001 | 0.0001 | 0.0001 | 0.0001 | 0.0001 |
| Mature cataract | Poor | 15 (8.0) | 91.4 (86.5-95.2) | 0.17 (-0.03-0.50) | 13.3 (0.00-41.7) | 98.3 (95.3-99.5) | 40.0 (0.0-100.0) | 92.9 (88.1-96.6) |
|  | Acceptable | 55 (5.1) | 96.0 (94.7-97.2) | 0.59 (0.47-0.69) | 61.82 (48.2-73.9) | 97.86 (96.7-98.6) | 60.7 (46.4-71.4) | 98.0 (96.9-98.8) |
|  | Optimal | 79 (9.3) | 96.0 (94.3-97.2) | 0.78 (0.70-0.85) | 89.9 (79.7-95.7) | 96.6 (94.8-97.8) | 73.2 (62.0-81.5) | 98.9 (97.8-99.6) |
|  | P value^e^ |  | 0.0001 | 0.0001 | 0.0001 | 0.0001 | 0.0001 | 0.0001 |
| Immature cataract | Poor | 110 (58.8) | 74.3 (66.8-81.3) | 0.49 (0.35-0.62) | 70.0 (60.5-79.4) | 80.5 (68.5-90.0) | 83.7 (73.9-92.0) | 65.3 (53.1-74.8) |
|  | Acceptable | 606 (56.0) | 84.0 (81.4-86.3) | 0.68 (0.63-0.73) | 78.1 (73.5-81.5) | 91.6 (88.6-94.2) | 92.2 (89.5-94.6) | 76.7 (72.6-80.3) |
|  | Optimal | 412 (48.7) | 87.4 (84.3-89.8) | 0.75 (0.69-0.80) | 78.6 (73.4-82.9) | 95.6 (92.5-97.6) | 94.5 (90.6-97.0) | 82.5 (78.1-86.1) |
|  | P value^e^ |  | 0.0001 | 0.0001 | 0.0001 | 0.0001 | 0.0001 | 0.0001 |
| Clear crystalline lens | Poor | 28 (15.0) | 81.3 (74.3-87.5) | 0.47 (0.29-0.63) | 85.7 (57.9-100.0) | 80.5 (73.1-87.4) | 43.6 (28.1-59.1) | 97.0 (89.8-100.0) |
|  | Acceptable | 190 (17.5) | 88.6 (86.3-90.7) | 0.66 (0.60-0.73) | 89.0 (82.9-93.5) | 88.6 (86.0-90.9) | 62.4 (55.3-69.9) | 97.4 (95.8-98.5) |
|  | Optimal | 120 (14.2) | 91.0 (88.4-93.3) | 0.68 (0.59-0.76) | 86.7 (77.4-92.8) | 91.7 (88.9-94.1) | 63.4 (53.8-72.6) | 97.7 (95.8-98.7) |
|  | P value^e^ |  | 0.0001 | 0.0001 | 0.0001 | 0.0001 | 0.0001 | 0.0001 |
| Pseudophakia | Poor | 33 (17.7) | 96.8 (93.0-99.0) | 0.89 (0.75-0.97) | 93.9 (78.6-100.0) | 97.4 (92.9-99.4) | 88.6 (70.2-97.4) | 98.7 (95.7-100.0) |
|  | Acceptable | 230 (21.2) | 96.9 (95.6-97.9) | 0.91 (0.87-0.94) | 94.8 (90.8-97.3) | 97.4 (96.0-98.4) | 90.8 (86.0-94.3) | 98.6 (97.5-99.3) |
|  | Optimal | 233 (27.5) | 97.8 (96.4-98.7) | 0.94 (0.91-0.97) | 97.4 (93.7-99.1) | 97.9 (96.5-98.9) | 94.6 (90.9-97.1) | 99.0 (97.5-99.7) |
|  | P value^e^ |  | 0.3924 (adjusted p=0.672) | 0.0001 | 0.0006 | 0.8924  (adjusted p=0.892) | 0.0001 | 0.0001 |
| Refractive error/ Presbyopia | Poor | 13 (7.0) | 83.4 (76.3-89.2) | -0.03 (-0.10-0.26) | 7.7 (0.0-50.0) | 89.1 (81.6-93.6) | 5.0 (0.0-31.3) | 92.8 (86.4-96.6) |
|  | Acceptable | 106 (9.8) | 88.1 (85.7-90.3) | 0.42 (0.31-0.52) | 57.6 (44.1-69.0) | 91.4 (89.2-93.4) | 42.1 (31.1-52.1) | 95.2 (93.3-96.7) |
|  | Optimal | 83 (9.8) | 87.1 (84.2-89.8) | 0.31 (0.20-0.45) | 41.0 (28.4-56.4) | 92.1 (89.7-94.4) | 36.2 (25.6-51.1) | 93.5 (91.3-95.6) |
|  | P value^e^ |  | 0.0001 | 0.0001 | 0.0001 | 0.0001 | 0.0001 | 0.0001 |

^a^ All values are shown as estimate (95% confidence interval) except where otherwise indicated.

^b^ Diagnosis was assigned by in-person eye camp ophthalmologist. “Any cataract” category combines both mature and immature cataract. Reference group for “cataract” is any other lens status including clear crystalline lens, pseudophakia, or aphakia. Pterygium, active corneal infection, and inactive corneal opacity were not analyzed due to small sample size (N<10) in at least one of the image quality subgroups.

^c^ Number (percentage) of eyes in each image quality group, as graded by remote ophthalmologists reviewing smartphone images.

^d^ Cohen’s kappa statistic is a measure of agreement and is evaluated as 0 (no agreement), 0.10-0.20 (poor agreement), 0.21-0.40 (fair agreement), 0.41-0.60 (moderate agreement), 0.61-0.80 (substantial agreement), 0.81-0.99 (near-perfect agreement), 1.0 (perfect agreement).

^e^ All P values were adjusted using the Benjamini–Hochberg procedure. However, only those explicitly labeled as “adjusted p” reflect this correction. For P values not labeled as adjusted, the Hochberg-adjusted values were identical to P values from the original non-parametric test since the values were <0.001.

Abbreviations: N = number

#### **Supplementary Table 3. Measures of diagnostic agreement between eye camp ophthalmologist in-person examination and remote ophthalmologist evaluation of smartphone-based screening data, by patient sex^a^**

| **Diagnosis^b^** | **Image Quality^c^** | **N (%)^c^** | **Percent agreement** | **Kappa statistic^d^** | **Sensitivity** | **Specificity** | **Positive predictive value** | **Negative predictive value** |
| --- | --- | --- | --- | --- | --- | --- | --- | --- |
| Any cataract | Male | 554 (62.9) | 87.2 (84.3-90.1) | 0.74 (0.68-0.80) | 82.0 (77.8-86.0) | 96.0 (92.7-98.1) | 97.2 (95.0-98.7) | 75.9 (70.4-80.8) |
|  | Female | 723 (58.4) | 89.5 (87.4-91.4) | 0.79 (0.75-0.83) | 85.2 (81.8-88.4) | 95.5 (92.9-97.5) | 96.4 (94.2-97.9) | 82.2 (78.3-85.7) |
|  | P value | 0.0001 | 0.0001 | 0.0001 | 0.0001 | 0.0001 | 0.0001 | 0.0001 |
| Mature cataract ^a^ | Male | 64 (7.3) | 95.2 (93.5-96.5) | 0.67 (0.56-0.76) | 75.0 (59.7-85.5) | 96.8 (95.4-97.9) | 64.9 (53.3-75.8) | 98.0 (96.6-98.9) |
|  | Female | 85 (6.9) | 95.9 (94.5-97.0) | 0.68 (0.59-0.77) | 69.4 (58.0-81.0) | 97.8 (96.8-98.6) | 70.2 (58.3-80.2) | 97.8 (96.6-98.6) |
|  | P value | 0.0001 | 0.0001 | 0.0001 | 0.0001 | 0.0001 | 0.0001 | 0.0001 |
| Immature cataract | Male | 490 (55.6) | 83.1 (80.1-86.1) | 0.67 (0.61-0.72) | 74.9 (70.5-79.8) | 93.4 (89.8-95.7) | 93.4 (90.0-95.8) | 74.8 (70.2-79.1) |
|  | Female | 638 (51.5) | 85.6 (83.1-87.7) | 0.71 (0.66-0.76) | 79.5 (75.5-83.0) | 92.0 (89.0-94.3) | 91.4 (88.1-94.0) | 80.9 (77.2-84.0) |
|  | P value | 0.0001 | 0.0001 | 0.0001 | 0.0001 | 0.0001 | 0.0001 | <0.0001 |
| Clear crystalline lens | Male | 135 (15.3) | 87.7 (84.7-90.4) | 0.61 (0.52-0.69) | 85.9 (76.8-91.6) | 88.1 (84.9-91.0) | 56.6 (46.8-65.2) | 97.2 (95.3-98.4) |
|  | Female | 205 (16.6) | 89.8 (87.5-91.7) | 0.68 (0.61-0.74) | 89.3 (82.6-93.5) | 89.9 (87.4-92.1) | 63.5 (55.9-70.8) | 97.7 (96.1-98.6) |
|  | P value | 0.0001 | 0.0001 | 0.0001 | 0.0001 | 0.0001 | 0.0001 | 0.0001 |
| Pseudophakia | Male | 189 (21.5) | 96.0 (94.2-97.3) | 0.89 (0.84-0.92) | 95.2 (90.6-97.8) | 96.2 (94.4-97.7) | 87.4 (81.4-92.0) | 98.7 (97.4-99.4) |
|  | Female | 309 (24.9) | 98.0 (97.0-98.6) | 0.95 (0.92-0.96) | 96.1 (92.9-98.1) | 98.6 (97.8-99.2) | 95.8 (93.0-97.5) | 98.7 (97.6-99.4) |
|  | P value | 0.0001 | 0.0001 | 0.0001 | 0.0001 | 0.0001 | 0.0001 | 0.0001 |
| Pterygium | Male | 32 (3.6) | 96.8 (95.0-98.1) | 0.55 (0.36-0.72) | 56.3 (36.2-77.9) | 98.4 (96.9-99.2) | 56.3 (34.9-77.3) | 98.4 (97.1-99.3) |
|  | Female | 94 (7.6) | 92.5 (90.5-94.0) | 0.43 (0.31-0.54) | 44.7 (32.7-56.4) | 96.4 (95.0-97.4) | 50.6 (36.8-63.1) | 95.5 (93.8-96.8) |
|  | P value | 0.0001 | 0.0001 | 0.0001 | 0.0001 | 0.0001 | 0.0001 | 0.0001 |
| Refractive error/ Presbyopia | Male | 107 (12.2) | 87.0 (83.8-89.6) | 0.39 (0.27-0.51) | 45.8 (33.3-58.6) | 92.6 (90.0-94.9) | 46.2 (34.6-60.6) | 92.5 (89.7-94.6) |
|  | Female | 95 (7.7) | 87.3 (84.7-89.6) | 0.31 (0.21-0.42) | 49.5 (36.0-62.8) | 90.4 (87.9-92.3) | 29.9 (21.2-40.0) | 95.6 (93.8-96.9) |
|  | P value | <0.0001 | <0.0001 | <0.0001 | 0.0001 | 0.0001 | 0.0001 | 0.0001 |

^a^ All values are shown as estimate (95% confidence interval) except where otherwise indicated.

^b^ Diagnosis was assigned by in-person eye camp ophthalmologist. “Any cataract” category combines both mature and immature cataract. Reference group for “cataract” is any other lens status including clear crystalline lens, pseudophakia, or aphakia. Pterygium, active corneal infection, and inactive corneal opacity were not analyzed due to small sample size (N<10) in at least one of the image quality subgroups.

^c^ Number (percentage) of eyes in each image quality group, as graded by remote ophthalmologists reviewing smartphone images.

^d^ Cohen’s kappa statistic is a measure of agreement and is evaluated as 0 (no agreement), 0.10-0.20 (poor agreement), 0.21-0.40 (fair agreement), 0.41-0.60 (moderate agreement), 0.61-0.80 (substantial agreement), 0.81-0.99 (near-perfect agreement), 1.0 (perfect agreement).

Abbreviations: N = number

#### **Supplementary Table 4. Odds of diagnostic concordance between remote ophthalmologists and in-person eye camp ophthalmologist, by sex**

| **Diagnosis^a^** | **Sex** | **N (%)** | **Odds Ratio (95% CI)^b^** | **P value** | **Adjusted P value** |
| --- | --- | --- | --- | --- | --- |
| Any cataract | Male | 557 (61.8) | 0.80 (0.6-1.1) | 0.186 | 0.472 |
|  | Female | 724 (57.6) | Ref | - | - |
| Mature cataract | Male | 64 (7.1) | 0.9 (0.5-1.4) | 0.511 | 0.767 |
|  | Female | 86 (6.9) | Ref | - | - |
| Immature Cataract | Male | 493 (54.7) | 0.8 (0.6-1.1) | 0.197 | 0.472 |
|  | Female | 638 (50.8) | Ref | - | - |
| Clear crystalline lens | Male | 135 (15.0) | 0.8 (0.6-1.2) | 0.236 | 0.472 |
|  | Female | 205 (16.3) | Ref | - | - |
| Pseudophakia | Male | 190 (21.1) | 0.5 (0.3-0.9) ^*^ | 0.020 ^*^ | 0.12 |
|  | Female | 309 (24.6) | Ref | - | - |
| Pterygium | Male | 34 (3.8) | 2.4 (1.4-4.1) ^*^ | 0.001 ^*^ | 0.012 ^*^ |
|  | Female | 97 (7.7) | Ref | - | - |
| Refractive error/presbyopia | Male | 118 (13.1) | 1.0 (0.7-1.4) | 0.869 | 0.892 |
|  | Female | 103 (8.2) | Ref | - | - |

^a^ Diagnosis was assigned by in-person eye camp ophthalmologist. “Any cataract” category combines both mature and immature cataract. Reference group for “cataract” is any other lens status including clear crystalline lens, pseudophakia, or aphakia. Pterygium, active corneal infection, and inactive corneal opacity were not analyzed due to small sample size (N<10) in at least one of the image quality subgroups.

^b^ Odds ratios estimated using univariable generalized estimating equations with a binomial distribution, logit link function, and robust standard errors assuming an exchangeable correlation structure at the patient level

Abbreviations: CI = confidence interval; N = number; Ref = reference group

### **SUPPLEMENTARY FIGURES**

#### **Supplementary Figure 1. InSightful mobile and web app used by CHWs for screening**

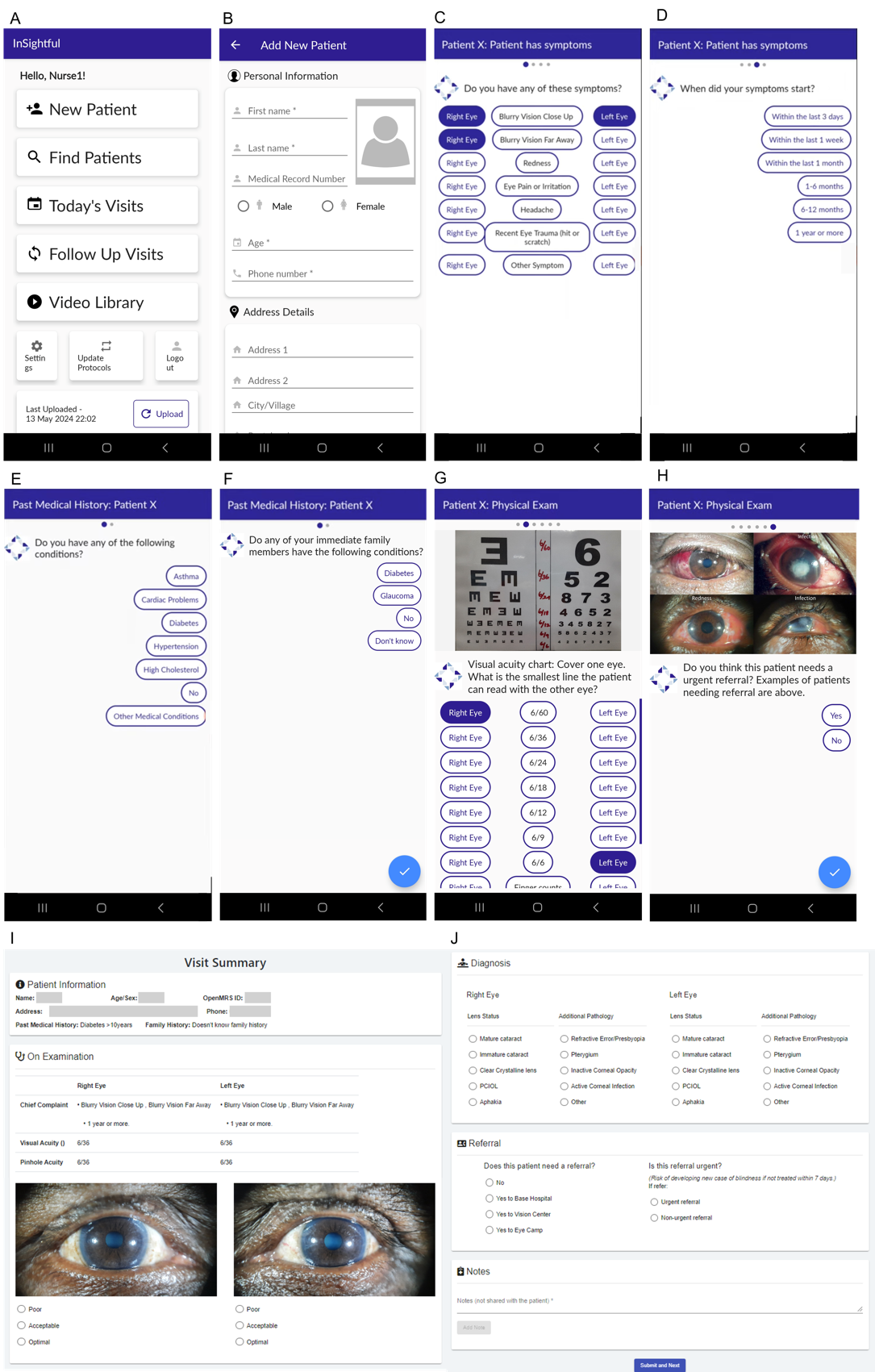
